# Development of a Rapid and Specific MALDI-TOF Mass Spectrometric Assay for SARS-CoV-2 Detection

**DOI:** 10.1101/2023.03.10.23287091

**Authors:** Lydia Kollhoff, Marc Kipping, Manfred Rauh, Uta Ceglarek, Günes Barka, Frederik Barka, Andrea Sinz

**Affiliations:** Department of Pharmaceutical Chemistry and Bioanalytics, Martin Luther University Halle-Wittenberg, Halle (Saale), Germany; Center for Structural Mass Spectrometry, Martin Luther University Halle-Wittenberg, Halle (Saale), Germany; Department of Pediatrics and Adolescent Medicine, Friedrich Alexander University Erlangen-Nürnberg, Germany; Institute for Laboratory Medicine, Clinical Chemistry and Molecular Diagnostics, University of Leipzig, Leipzig, Germany; SunChrom Wissenschaftliche Geräte GmbH, 61381 Friedrichsdorf, Germany

**Keywords:** COVID-19, MALDI, Mass spectrometry, SARS-COV-2

## Abstract

We have developed a rapid and highly specific assay for detecting and monitoring SARS-CoV-2 infections by matrix-assisted laser desorption/ionization time-of-flight mass spectrometry (MALDI-TOF-MS). As MALDI-TOF mass spectrometers are available in a clinical setting, our assay has the potential to serve as alternative to the commonly used reverse transcriptase quantitative polymerase chain reaction (RT-qPCR). Sample preparation prior to MALDI-TOF-MS involves the tryptic digestion of SARS-CoV-2 proteins, followed by an enrichment of virus-specific peptides from SARS-CoV-2 nucleoprotein via magnetic antibody beads. Our MALDI-TOF-MS method allows the detection of SARS-CoV-2 nucleoprotein as low as 8 amol/μl. MALDI-TOF mass spectra are obtained in just a few seconds, which makes our MS-based assay suitable for a high-throughput screening of SARS-CoV-2 in healthcare facilities in addition to PCR. Due to the specific detection of virus peptides, different SARS-CoV-2 variants are readily distinguished from each other. Specifically, we show that our MALDI-TOF-MS assay discriminates SARS-CoV-2 strain B.1.617.2 “delta variant” from all other variants in patients’ samples, making our method highly valuable to monitor the emergence of new virus variants.

**GRAPHICAL ABSTRACT:** 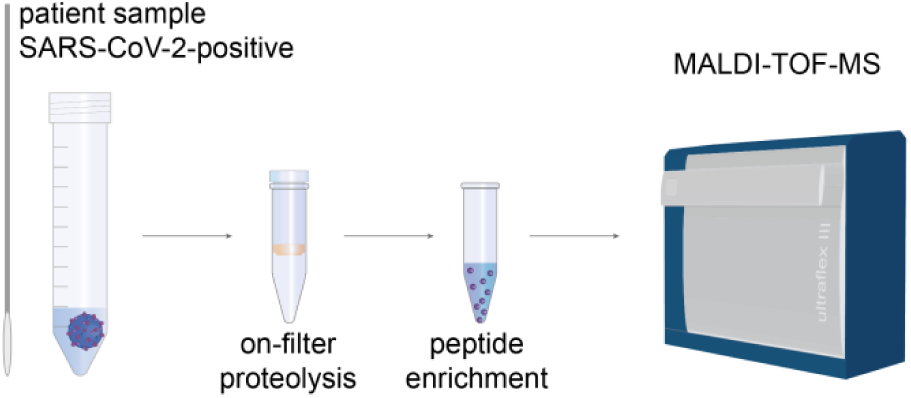

## INTRODUCTION

The COVID-19 pandemic is still ongoing with a total of 753 million cases worldwide so far^1^. The surveillance of new infections, especially in clinics and other healthcare facilities is essential not only for preventing infections of vulnerable groups, but also for monitoring the emergence and spreading of new virus variants.

Reverse transcriptase quantitative polymerase chain reaction (RT-qPCR) has been the method-of-choice for detecting and monitoring severe acute respiratory syndrome coronavirus 2 (SARS-CoV-2) infections since the beginning of the pandemic. The main advantage of RT-qPCR is its high sensitivity, but as it relies on amplifying the genetic material of the virus, the PCR method also has its limitations. The concentration of viral RNA in a sample can be estimated, but the exact amount of virus particles cannot be determined by the cycle time (Ct value) of a sample alone^2^. Also, patients who recover from an infection might still be identified as SARS-CoV-2-positive by PCR, despite the lack of infectious virus particles in the sample^3^.

Therefore, alternative methods to RT-qPCR are needed providing a more reliable quantitative determination of virus particles in a sample for an accurate detection and monitoring of SARS-CoV-2 infections. Mass spectrometric (MS) techniques can provide qualitative as well as quantitative information of virus-specific proteins with high accuracy and precision^4^. Numerous MS-based methods have been described for the detection of SARS-CoV-2 since the outbreak of the pandemic. Most of the methods rely on the detection of viral peptides^6,8,7,5^. By measuring the total amount of viral peptides instead of amplified RNA, virus amounts are directly determined and different virus variants can be identified in the samples ^8,7^. This makes MS a highly valuable tool for detecting and monitoring the presence of specific virus variants as well as the emergence of new variants. The inherent advantages of MS-based assays to detect SARS-CoV-2 infections are their high sensitivity, their speed, the potential to use them in a routine clinical environment, and their cost efficiency per sample.

Following our previous results of developing a specific SARS-CoV-2 detection method based on liquid chromatography tandem mass spectrometry (LC-MS/MS)^9,10^, we now introduce a method for SARS-CoV-2 detection by matrix-assisted laser desorption/ionization time-of-flight mass spectrometers (MALDI-TOF-MS). MALDI-TOF mass spectrometers are available in a lot of clinical laboratories and are used for diagnosing bacterial, fungal, and viral infections^11^. With MALDI-TOF-MS, a sample can be measured within seconds, making sample preparation the limiting factor in terms of high-throughput analysis. By automating and parallelizing sample preparation, results from our MS assay are obtained in less than two hours, even for large sample numbers.

## MATERIALS AND METHODS

### Chemicals and Materials

All chemicals were obtained from Sigma Aldrich (Taufkirchen, Germany) at the highest purity available. Cobas PCR Medium was obtained from Roche Diagnostics GmbH (Mannheim, Germany). Enzymes were obtained from Promega GmbH (Mannheim, Germany). Magnetic antibody beads against peptides of the SARS-CoV-2 nucleoprotein listed in **Table 1** were obtained from SISCAPA Assay Technologies (Washington, DC). 300-kDa Molecular weight cut-off (MWCO) filters (Nanosep) were obtained from Pall Filtersystems GmbH (Bad Kreuznach, Germany). Recombinant SARS-CoV-2 nucleoprotein was obtained from antibodies-online GmbH (Aachen, Germany). Stable isotope-labeled peptides (SpikeTide-TQL) derived from SARS-CoV-2 nucleoprotein were obtained from JPT Peptide Technologies GmbH (Berlin Germany). The SARS-CoV-2 LC-MS Kit (RUO) was obtained from Waters (Eschborn, Germany).

**Table 1.**
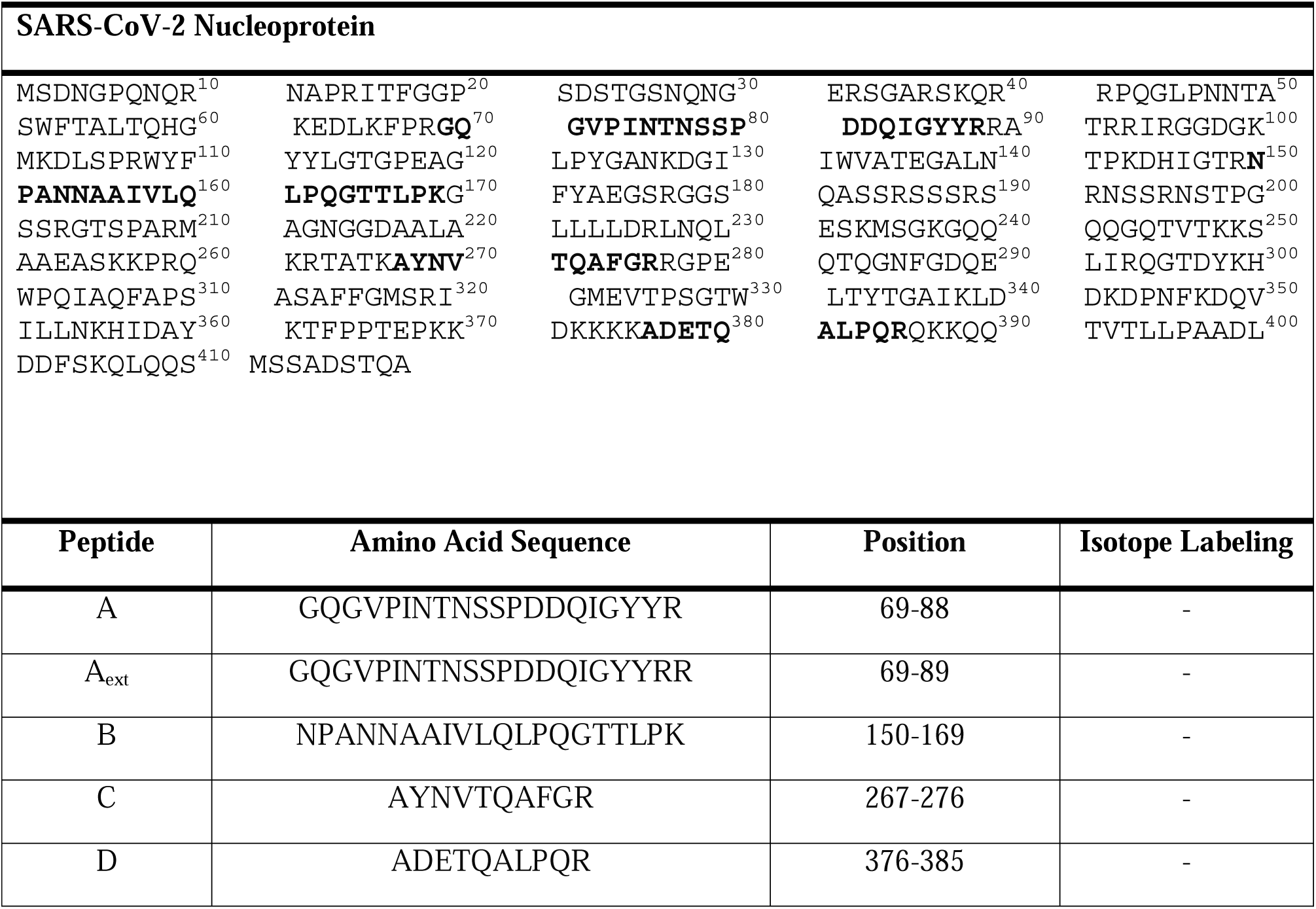
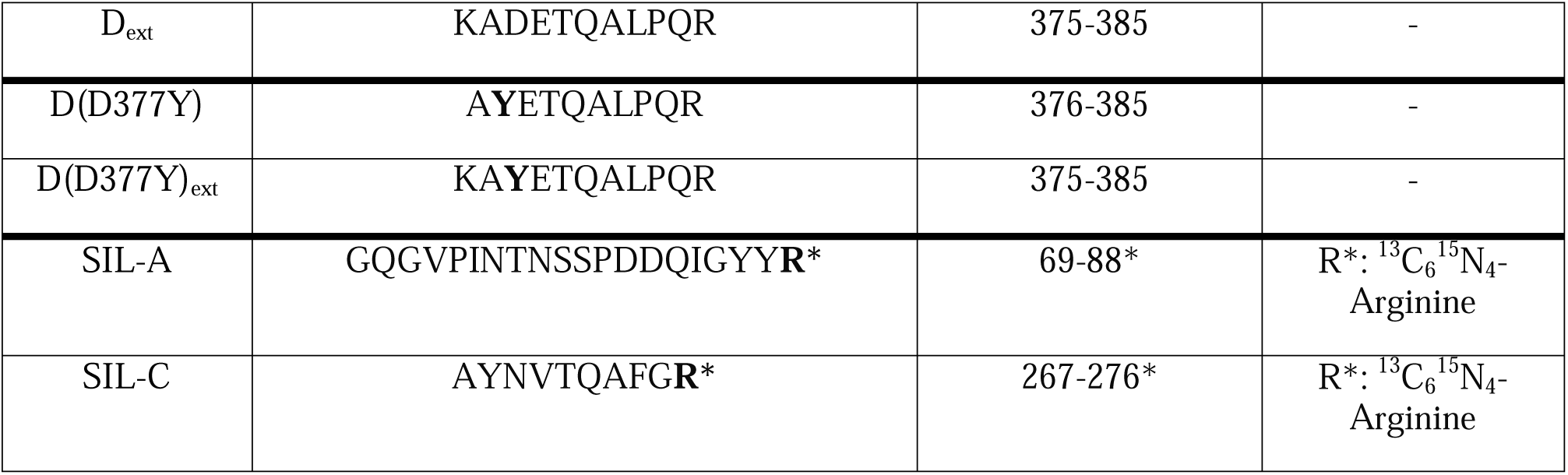
Amino Acid Sequences of SARS-CoV-2 Nucleoprotein (Upper Panel) and Tryptic Peptides (Lower Panel). Amino acids corresponding to peptides (A-D) that were used for the MALDI-TOF-MS assay are printed in bold. Peptide D was detected in two variants (D377 and Y377). Peptides A and D were additionally used in their extended forms (ext) with one missed cleavage site of trypsin; peptides A and C were also employed as stable isotope-labeled peptides (SIL) with C-terminal arginine residues labeled (marked as R*).

### Sample Collection

Nasopharyngeal swab samples were provided by the University Clinics Erlangen, Germany. Samples were collected in 600 μl Cobas PCR Medium and 300 μl were used for RT-qPCR analysis. The rest of the sample was heat-inactivated (3 h, 70°C) and stored at 4 °C.

### Sample Preparation for Method Development

For method development, recombinant SARS-CoV-2 nucleoprotein was spiked into nasopharyngeal swab samples of 30 healthy volunteers. Here, the recombinant protein was added after the filtration step as explained in the next paragraph.

600 μl of nasopharyngeal swab samples from healthy volunteers were loaded on 300-kDa MWCO filters respectively, centrifuged at 5,000 g for 5 minutes and washed twice with 200 μl of 100 mM ammonium bicarbonate. Recombinant SARS-CoV-2 nucleoprotein was spiked at concentrations from 22,5 amol/μl to 450 amol/μl into 100 μl of the samples on the filter prior to enzymatic proteolysis. The concentrations of SARS-CoV-2 nucleoprotein correspond to theoretical concentrations of 3.75 amol/μl to 75 amol/μl in 600 μl of nasopharyngeal swab sample.

### Patient Sample Preparation

300 μl nasopharyngeal swab samples of 53 SARS-CoV-2-positive individuals were loaded on 300-kDa MWCO filters respectively, centrifuged at 5,000 g for 5 minutes and washed twice with 200 μl of 100 mM ammonium bicarbonate.

### Enzymatic Proteolysis

20 fmoles (20 μl of 1 fmol/μl solution in 100 mM ammonium bicarbonate) of isotope-labeled SIL-A and SIL-C peptides derived from SARS-CoV-2 nucleoprotein (**Table 1)** and 1 μg trypsin/Lys-C (1:1) protease mixture were added after filtration to either samples spiked with recombinant nucleoprotein or to SARS-CoV-2-positive patient samples.

Proteolysis was performed directly on the filter at 37 °C for 30 minutes. Digestion mixtures were centrifuged at 8,000 g for 5 minutes and collected in 2-ml-Eppendorf tubes. 300-kDa MWCO filters were washed with 20 μl of 0.5 M NaCl solution. Enzymatic proteolysis was stopped by adding 10 μg (20 μl) of tosyl-L-lysyl-chloromethane hydrochloride (TLCK) and incubating the solution at 1,000 rpm (5 min, room temperature).

### Peptide Enrichment Prior to MALDI-TOF-MS

For peptide enrichment, monoclonal antibodies specific for tryptic peptides of SARS-CoV-2 nucleoprotein A and C or A and D (**Table 1**), and coupled to magnetic beads, were added to the samples. The solutions were incubated at 1,400 rpm for 1 h at room temperature. After incubation, samples were placed on a magnetic rack and beads were washed twice with 150 μl of phosphate buffered saline buffer (PBS, pH 7.4). Antibody-bound peptides were eluted with 2 μl of 1% (v/v) trifluoroacetic acid (TFA).

Prior to MALDI-TOF-MS analysis, 1 μl of elution solutions were mixed with 1 μl of matrix solution (0.7 mg/ml α-cyano-4-hydroxycinnamic acid (CHCA) in 40 % (v/v) acetonitrile/ 0.1 % (v/v) TFA). 1 μl of this mixture was spotted on an AnchorChip target (Bruker Daltonik, Bremen, Germany).

### MALDI-TOF-MS

Mass spectra were obtained with an Ultraflex III MALDI-TOF/TOF mass spectrometer (Bruker Daltonik, Bremen). The instrument was operated in positive ionization and reflectron mode. The mass range was set to 600-5000 Da. For each sample, 1000 spectra were accumulated. For external mass calibration, Peptide Calibration Mix II (Bruker Daltonik, Bremen) was employed.

### Peptide Enrichment Prior to LC-ESI-MS/MS

For LC-ESI-MS/MS preparation, the SARS-CoV-2 LC-MS Kit was used. Monoclonal antibodies specific for tryptic peptides of SARS-CoV-2 nucleoprotein (**Table 1**) that had been coupled to magnetic beads, were added to the samples and the solutions were incubated as described above. After incubation, samples were placed on a magnetic rack and beads were washed twice with 150 μl of CHAPS (3-((3-cholamidopropyl) dimethylammonio)-1-propanesulfonate) washing buffer (0.03 % (w/v) CHAPS in PBS buffer, pH 7.4). Enriched SARS-CoV2 peptides were eluted with 50 μl of CHAPS elution buffer (0.03 % (w/v) CHAPS in 1 % (v/v) formic acid).

### LC-ESI-MS/MS

LC separation of peptides was performed on a UPLC I-Class FTN system (Waters, Eschborn, Germany) equipped with a BEH C18 column (2.1 mm x 50 mm, 1.7 μm, Waters). The UPLC system was directly coupled to a Xevo TQ-XS mass spectrometer (Waters, Eschborn, Germany) equipped with ESI source. MS acquisition was performed using a multiple-reaction monitoring (MRM) method of three selected transitions for each SARS-CoV-2 nucleoprotein peptide B, C, and D (**Supporting Information, Table S1**).

## RESULTS AND DISCUSSION

### Development of SARS-CoV-2 Assay

The workflow of our MALDI-TOF-MS assay to detect SARS-CoV-2 is summarized in **Figure 1**. Emphasis was laid on optimizing the sensitivity for detecting specific peptides derived from SARS-CoV-2 nucleoprotein (**Table 1**). To remove buffer components and reduce sample complexity, a filter-aided sample preparation step is required prior to enzymatic proteolysis. With a molecular weight cut-off of 300 kDa, viral particles remain on the filter, while buffer components and soluble proteins pass through the filter^12^. Enzymatic proteolysis of proteins in the sample was then performed directly on the filter.

**Figure 1.**
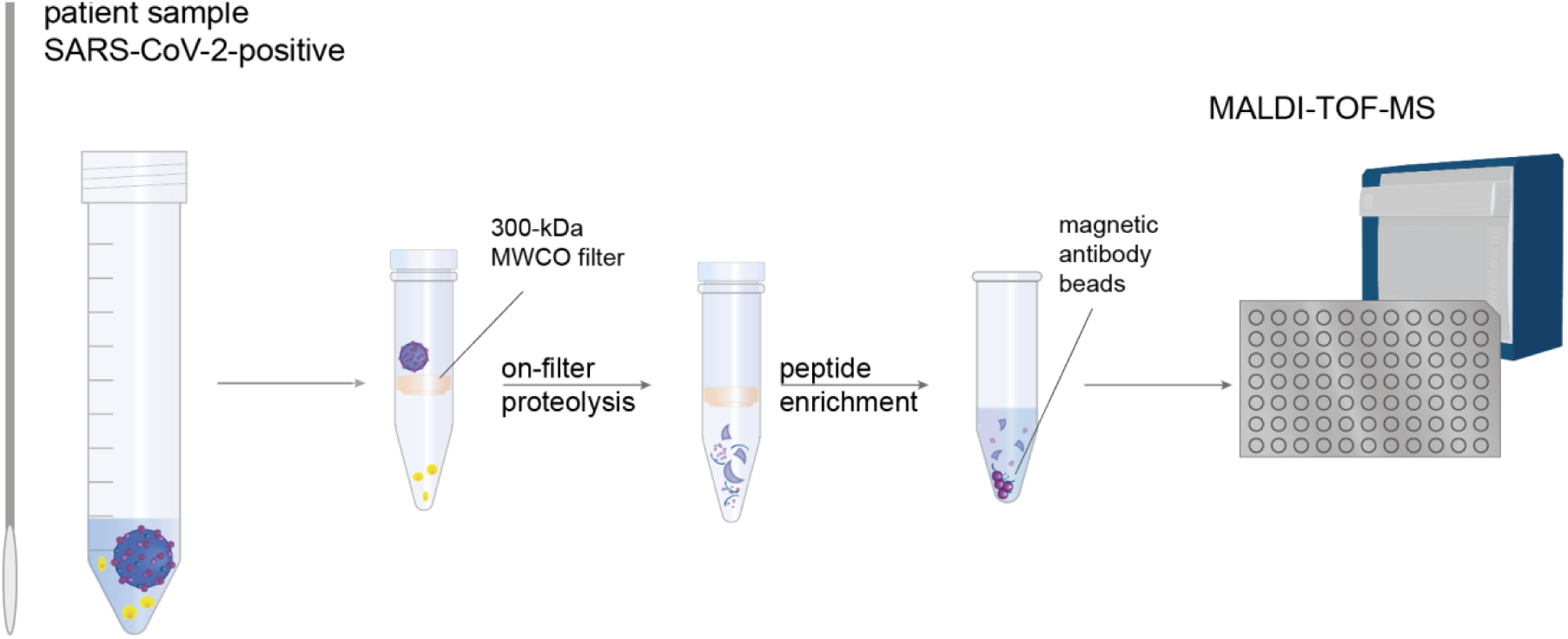
Analytical Workflow of MALDI-TOF-MS Assay for SARS-CoV-2 Detection. The method is based on a specific detection of peptides derived from SARS-CoV-2 nucleoprotein. Nasopharyngeal swabs are collected and loaded on a 300-kDa molecular weight cut-off filter. After enzymatic proteolysis, specific peptides of SARS-CoV-2 nucleoprotein are enriched with magnetic antibody beads and analyzed by MALDI-TOF-MS.

The application of magnetic antibody beads for a specific enrichment of peptides derived from SARS-CoV-2 nucleoprotein after tryptic digestion has already been successfully demonstrated for LC-MS/MS workflows^13 14^. Magnetic antibody beads are available for different tryptic peptides of the SARS-CoV-2 nucleoprotein, but enriched peptides might exhibit different ionization efficiencies in ESI- and MALDI-MS. To test the applicability of magnetic antibody beads directed against SARS-CoV-2 nucleoprotein in a MALDI-TOF-MS workflow, recombinant SARS-CoV-2 nucleoprotein was enzymatically digested and analyzed by MALDI-TOF-MS. Peptide C (AYNVTQAFGR, see **Table 1**) from SARS-CoV-2 nucleoprotein was found to exhibit one of the most intense signals in the mass spectrum (Supporting Information, **Figure S1**), followed by peptide A (GQGVPINTNSSPDDQIGYYR). The signal of peptide D (ADETQALPQR, [M+H]^+^ at *m/z* 1128.6) of SARS-CoV-2 nucleoprotein overlaps with the signal of peptide C ([M+H]^+^ at *m/z* 1126.6), (Supporting Information, **Figure S2)**. An antibody enrichment of both peptides C and D results in signal suppression of peptide D, hampering an accurate quantification due to overlapping signals. Conclusively, the enrichment of peptides A and C from SARS-CoV-2 nucleoprotein (**Table 1**) allows a specific and sensitive virus detection by MALDI-TOF-MS. One of the most important advantages of MALDI-TOF-MS is its speed with analysis times of a few seconds per sample. Calculating 30 minutes for proteolytic digestion and one hour for peptide enrichment, a sample batch can be prepared and measured in less than two hours. This makes our MALDI-TOF-MS assay a highly appealing option for high-throughput applications in a clinical setting.

### Quantification of SARS-CoV-2 Peptides

For virus quantification, stable isotope-labeled tryptic peptides of SARS-CoV-2 nucleoprotein were employed (see **Table 1**). As outlined above, peptides A and C exhibit the most prominent signals in MALDI-TOF-MS, therefore quantification might be performed using their isotope-labeled peptide counterparts SIL-A or SIL-C. We selected SIL-C as optimal candidate for quantification as its signal intensities are usually higher than for SIL-A (Supporting Information, **Figures S3 and S4** and **Figure 2A** for SARS-CoV-2-positive patient samples). Also, the standard deviations of signal intensities obtained with peptide SIL-A in repeated MALDI-TOF-MS measurements are higher than for SIL-C, especially at higher concentrations (Supporting Information, **Figure S4**). Peptide SIL-A might however still serve as confirmation for SARS-CoV-2-positive samples. Quantification with SIL-C was optimized for SARS-CoV-2 nucleoprotein concentrations between ∼50 to 600 amol/μl (Supporting Information, Figure S5).

**Figure 2.**
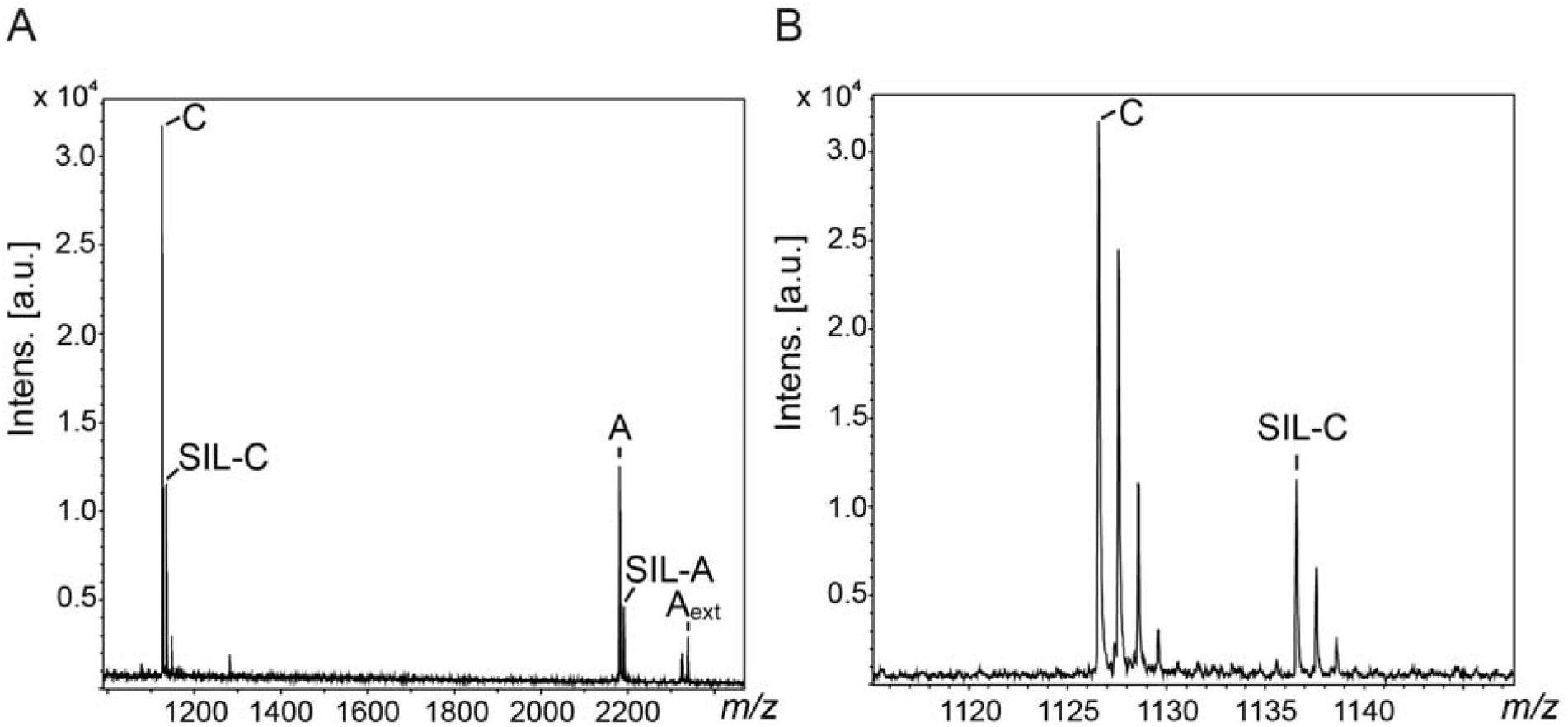
MALDI-TOF-Mass Spectra of SARS-CoV-2-Positive Patient Samples. (A) Tryptic peptides from SARS-CoV-2 nucleoprotein GQGVPINTNSSPDDQIGYYR (peptide A), GQGVPINTNSSPDDQIGYYRR (peptide A_ext_) and AYNVTQAFGR (peptide C) as well as the isotope-labeled peptides SIL-A and SIL-C (**Table 1**) are highlighted. **(B)** Diagnostic peptides from SARS-CoV-2 nucleoprotein (peptides C and SIL-C) are shown enlarged.

### Sensitivity and Comparability of MALDI-TOF-MS ASSAY Compared to RT-qPCR

The sensitivity of our MALDI-TOF-MS assay was determined by spiking recombinant SARS-CoV-2 nucleoprotein into nasopharyngeal samples from healthy individuals. Nucleoprotein was spiked into the samples at different concentrations after centrifugation. Due to its molecular weight of 45.6 kDa it can pass through the 300-kDa MWCO filter.

With a variation coefficient <25 % and a bias of ± 20 % in 6 biological replicates, the limit of quantitation (LOQ) was determined to be 8 amol/μl, corresponding to a RT-qPCR cycle time (Ct) value of 24^15^. Our MALDI-TOF-MS assay does not fully reach the sensitivity of RT-qPCR, but samples with a high viral load will be detected. Therefore, infective patients might rapidly be identified in large cohorts, which is highly valuable for detecting and monitoring virus outbreaks and will help in disrupting infection chains earlier.

### Application of the MALDI-TOF-MS Assay to SARS-CoV-2-Positive Patient Samples

After having developed a stable and reproducible workflow for identifying peptides of spiked-in SARS-CoV-2 nucleoprotein from nasopharyngeal samples, we sought to apply our MALDI-TOF-MS assay for detecting viral peptides in confirmed SARS-CoV-2-positive patient samples. **Figure 2** shows mass spectra of a SARS-CoV-2-positive sample with diagnostic peptides A and C, together with the externally added isotope-labeled counterparts SIL-A and SIL-C. Both peptides A and C ([M+H]^+^ at *m/z* 2181.0 and *m/z* 1126.6) yield prominent signals due to the specific antibody-based enrichment (**Figure 1**). In addition, peptide A appears with a missed cleavage site with [M+H]^+^ at *m/z* 2337.1 (A_ext_, see **Table 1**). The intense signals of enriched SARS-CoV-2 peptides allow a straightforward virus detection in the nasopharyngeal sample. As for the samples that had been spiked with recombinant SARS-CoV-2 nucleoprotein, peptide C yields higher signal intensities than peptide A. Peptide C allows quantifying the amount of nucleoprotein in the patient samples, while peptide A serves as confirmation of a SARS-CoV-2 infection.

Using the 53 SARS-CoV-2-positive samples with known Ct values **(Figure 3**), we were able compare the outcome of our MALDI-TOF-MS assay with RT-qPCR results. SARS-CoV-2 nucleoprotein can be quantified with our MS-method up to Ct value of 22, which is lower than the LOQ obtained with the samples containing recombinant nucleoprotein (see above). This might be caused by a more complex matrix in SARS-CoV-2-positive patient samples or by loss of material during preparation as only intact viral particles remain on the filter for a subsequent enzymatic digestion. Sample loss during the filtration steps cannot be avoided, but it might be reduced to a minimum if centrifugation time and centrifugal force are reduced. Also, heat inactivation prior to sample preparation, which might also contribute to sample loss, is usually not required -especially when sample preparation workflows are fully automated in the clinical environment.

**Figure 3.**
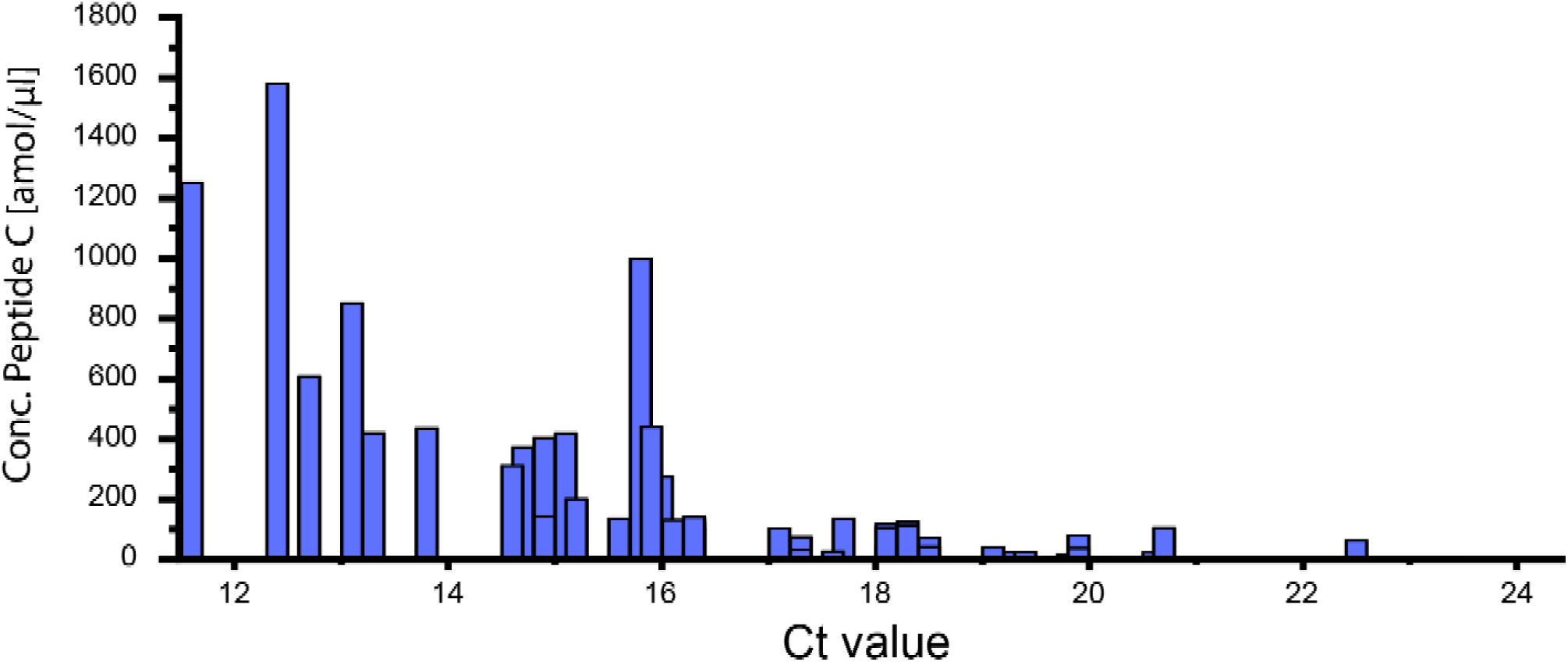
Overview of 53 SARS-CoV-2-Positive Nasopharyngeal Samples. Ct values in RT-qPCR are compared to the concentration of peptide C from SARS-CoV-2 nucleoprotein as determined by MALDI-TOF-MS in the same samples.

Interestingly, a number of patient samples with identical or similar Ct values were found to vary remarkably in their concentrations as determined with our MALDI-TOF-MS method. With the relative quantification of SARS-CoV-2 nucleoprotein based on isotope-labeled peptides, our assay directly reflects the viral concentration in a sample. RT-qPCR relies on the amplification of the virus’ genetic material, therefore Ct values will only give an indirect measure of the viral load. This issue has already been discussed previously.^2^ Also, residual viral RNA might still be present in recovered patients, which will be amplified as well and ultimately lead to false Ct values in RT-qPCR.^3^ It is important to note in this context that false positive SARS-CoV-2 detection is definitely ruled out in our assay as peptides from SARS-CoV-2 nucleoprotein are specifically enriched by the antibodies used herein and analysis is focused on virus-specific signals in MALDI-TOF-MS.

### Detection of SARS-CoV-2 Strain B.1.617.2 “Delta Variant”

In the course of method development, samples were initially analyzed by LC-MS/MS. For the LC-MS/MS workflow, peptides B, C and D from SARS-CoV-2 nucleoprotein were enriched on antibody magnetic beads, as outlined above. While the transitions for peptide B and C in LC-MS/MS multiple reaction analysis (MRM) were visible for all SARS-CoV-2 positive patient samples, we observed that in some samples, specific transitions for peptide D were missing in MRM LC-MS/MS analyses (see **Figure 4)**.

**Figure 4.**
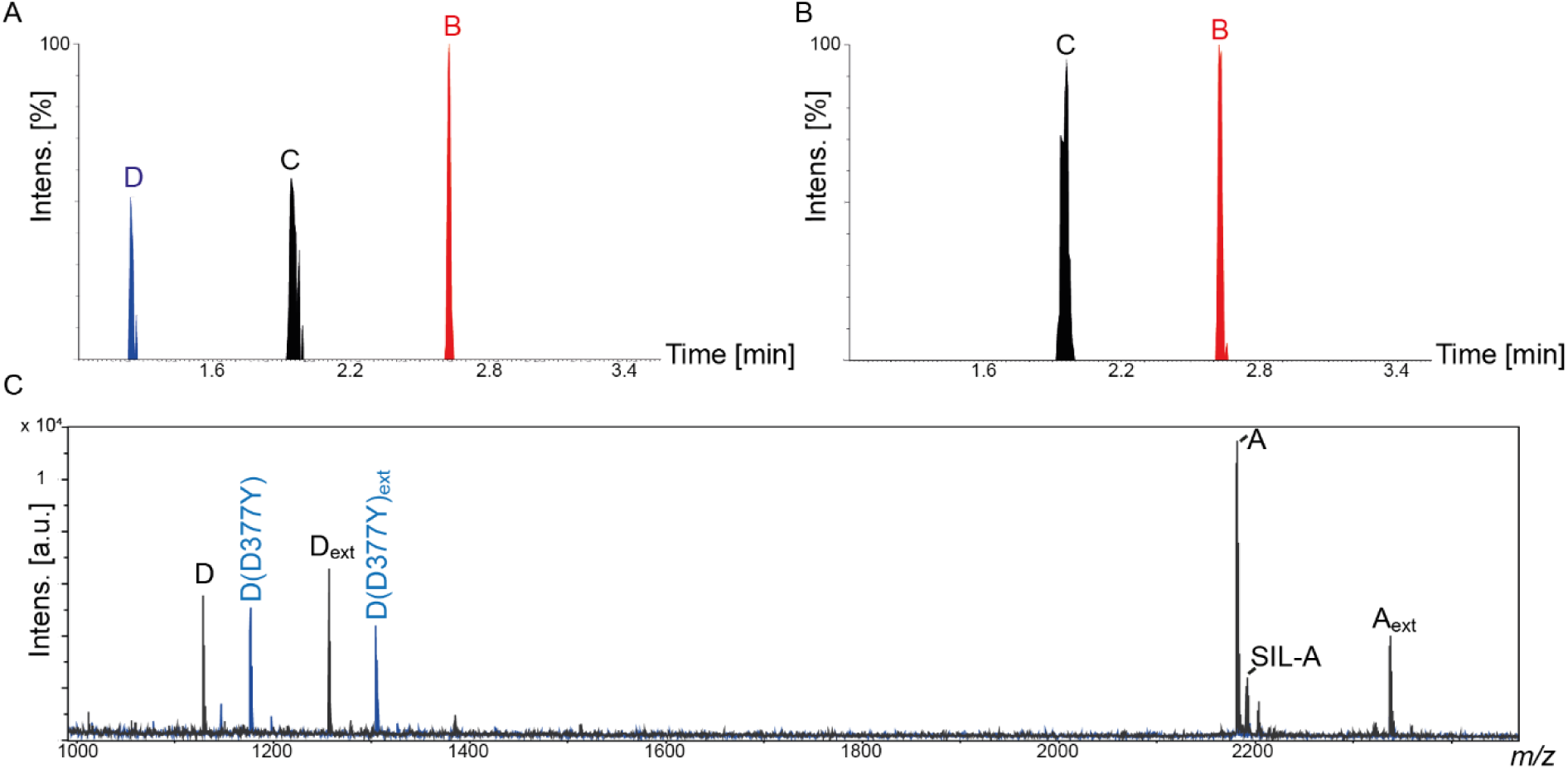
Discrimination of SARS-CoV-2 “Delta Variant” from other Viral Strains. (A) MRM chromatogram of a SARS-CoV-2-positive patient sample with selected transitions for peptide B (NPANNAAIVLQLPQGTTLPK), peptide C (AYNVTQAFGR, and peptide D (ADETQALPQR). (B) MRM chromatogram of a SARS-CoV-2-positive sample with selected transitions for peptides B and C. (C) MALDI-TOF mass spectra of SARS-CoV-2 strain B.1.1.529 “omicron variant” (black) and SARS-CoV-2 strain B.1.617.2 “delta variant” (blue) from nasopharyngeal samples. Peptide D (ADETQALPQR), peptide D_ext_ (KADETQALPQR), peptide D(D377Y) (AYETQALPQR), and peptide D(D377Y)_ext_ (KAYETQALPQR) are highlighted. Peptides A (GQGVPINTNSSPDDQIGYYR), A_ext_ (GQGVPINTNSSPDDQIGYYRR), and isotope-labeled peptide SIL-A (GQGVPINTNSSPDDQIGYYR*) are visible in both samples.

This interesting observation is easily explained by a mutation of peptide D in the SARS-CoV-2 strain delta B.1.617.2 “delta variant” at position 377 where the aspartic acid is replaced by a tyrosine residue^14^. All 53 SARS-CoV-2-positive nasopharyngeal patient samples were collected during December 2021 and January 2022. During this time, the “delta variant” (strain B.1.617.2) was successively replaced by the “omicron variant” (strain B.1.1.529) of SARS-CoV-2^16^ and the appearance of a new virus variant was successfully monitored by MS.

We then used our MALDI-TOF-MS assay to confirm our LC-MS/MS findings. For this purpose, antibody magnetic beads targeting peptides A and D were used for enrichment. Peptide C was not enriched as signals of peptides C and D would overlap in the MALDI-TOF mass spectra leading to signal suppression (see above).

In **Figure 4 C**, the MALDI-TOF mass spectra of two patient samples are compared. Both samples has been classified by RT-qPCR as SARS-CoV-2-positive. In our MALDI-TOF-MS assay, high-intense signals of peptide D are detected in one sample, with and without tryptic missed cleavage sites (*m/z* 1128.6 for peptide D; *m/z* 1256.7 for peptide D_ext_). In another sample, these signals are missing, but instead two high-intense signals are visible at *m/z* 1176.6 and 1304.7. These signals were unambiguously assigned to versions of peptide D, in which the aspartic acid at position 377Y has been mutated to a tyrosine residue (D377Y). This mutation is a clear indication of the SARS-CoV-2 “delta variant” and impressively shows the power of MALDI-TOF-MS assay that allows distinguishing different virus variants.

The SARS-CoV-2 nucleoprotein is less susceptible to mutations compared to spike protein, but mutations have been described for the nucleoprotein of several SARS-CoV-2 variants^17^. This makes peptides from the SARS-CoV-2 nucleoprotein a robust template for our MALDI-TOF-MS method as the antibody-based enrichment of specific peptides works for different viral strains. This underlines the strength of our assay as virus mutations will immediately be detected in the mass spectra and the method can be adapted within a few hours.

## CONCLUSIONS

We have developed a rapid and sensitive MS-based assay that allows a reliable detection of SARS-CoV-2 infections in patient samples. For an enrichment of peptides from SARS-CoV-2 nucleoprotein, a specific antibody enrichment was employed to increase the sensitivity of our assay. However, the sensitivity of current RT-qPCR methods is not yet met by our MALDI-TOF-MS assay, but it allows identifying patients with a high viral load corresponding to Ct values < 24 in RT-qPCR. The main advantage of our MS assay is the possibility to identify COVID-19 infections within a short time without identifying false positives. This might contribute to a fast control of COVID-19 outbreaks as test results are obtained in less than two hours for a large patient cohort by automating the sample preparation workflow. A reliable quantification of SARS-CoV-2 virus particles will also allow an accurate monitoring of the course of an infection in individual patients over time. The SARS-CoV-2 delta variant was distinguished from other viral variants of the SARS-CoV-2 virus in our MS assay. This will pave the way to rapidly detect upcoming new virus variants and to take appropriate measures to protect individuals from an infection.

## Data Availability

All data produced in the present work are contained in the manuscript.

## ASSOCIATED CONTENT

### Supporting Information

The Supporting Information is available free of charge on the ACS Publications website and contains the following:

**Table S1**. LC-MS/MS MRM (multiple reaction monitoring) method settings using the SARS-CoV-2 LC-MS Kit (RUO, Waters Corp).

**Figure S1**. MALDI-TOF mass spectrum of tryptic peptides from SARS-CoV-nucleoprotein.

**Figure S2**. MALDI-TOF mass spectrum of peptides C and D (antibody-enriched) from SARS-CoV-2 nucleoprotein.

**Figure S3**. MALDI-TOF mass spectrum of peptides A and C (antibody-enriched) from SARS-CoV-2 nucleoprotein, spiked into Cobas PCR Medium.

**Figure S4**. Comparison of SARS-CoV-2 quantification with peptides SIL-A and SIL-C.

**Figure S5**. Quantification of SARS-CoV-2 nucleoprotein with peptide SIL-C.

## AUTHOR INFORMATION

Address correspondence to

Corresponding author’s address: Department of Pharmaceutical Chemistry and Bioanalytics, Center for Structural Mass Spectrometry, Martin Luther University Halle-Wittenberg, Kurt-Mothes-Str. 3, D-06120 Halle (Saale), Germany.

## Author Contributions

LK, MK, and AS designed the experiments; LK analyzed the data and wrote the initial draft of the manuscript. MR and UC provided samples; FB and GB provided equipment. AS and LK revised the manuscript; AS secured funding. All authors have given approval to the final version of the manuscript.

## ACKNOWLEDGMENTS

This project is funded by the Federal Ministry for Economic Affairs and Energy (BMWi, ZIM project KK5096401SK0AS). AS acknowledges financial support by the DFG (RTG 2467, project number 391498659 “Intrinsically Disordered Proteins – Molecular Principles, Cellular Functions, and Diseases”, INST 271/404-1 FUGG, INST 271/405-1 FUGG, and CRC 1423, project number 421152132), the region of Saxony-Anhalt, and the Martin Luther University Halle-Wittenberg (Center for Structural Mass Spectrometry). The authors thank Matt Pope from SISCAPA for helpful discussions.

## Notes

### Competing Interest Statement

The authors have declared no competing interest.

### Funding Statement

This study was funded by the Federal Ministry for Economic Affairs and Energy (BMWi, ZIM project KK5096401SK0AS).

### Author Declarations

Ethics committee of the University Clinics Leipzig gave ethical approval for this work.

