## Supplemental Information for "Development of a Rapid and Specific MALDI-TOF Mass Spectrometric Assay for SARS-CoV-2 Detection"

**Table of Contents**

**Table S1**.	LC-MS/MS MRM (multiple reaction monitoring) method settings using the SARS-CoV-2 LC-MS Kit (RUO, Waters Corp).

**Figure S1.**	MALDI-TOF mass spectrum of tryptic peptides from SARS-CoV-nucleoprotein.

**Figure S2.**	MALDI-TOF mass spectrum of peptides C and D (antibody-enriched) from SARS-CoV-2 nucleoprotein.

**Figure S3.**	MALDI-TOF mass spectrum of peptides A and C (antibody-enriched) from SARS-CoV-2 nucleoprotein, spiked into Cobas PCR Medium.

**Figure S4.**	Comparison of SARS-CoV-2 quantification with peptides SIL-A and SIL-C.

**Figure S5.**	Quantification of SARS-CoV-2 nucleoprotein with peptide SIL-C.

**Table S1.** LC-MS/MS MRM (multiple reaction monitoring) method settings using the SARS-CoV-2 LC-MS Kit (RUO, Waters Corp).

| **Peptide** | **MRM** |  | **Cone (V)** | **Collison (V)** | **Retention Time (min)** | **Scan Window (min)** |
| --- | --- | --- | --- | --- | --- | --- |
| D | 564.8>400.2 | Quantifier | 35 | 19 | 0.55 | 0.40-0.70 |
|  | 564.8>584.4 | Qualifier | 35 | 20 | 0.55 | 0.40-0.70 |
|  | 564.8>712.4 | Qualifier | 35 | 24 | 0.55 | 0.40-0.70 |
| C | 563.8>679.4 | Quantifier | 35 | 19 | 0.90 | 0.71-1.1 |
|  | 563.8>578.3 | Qualifier | 35 | 18 | 0.90 | 0.71-1.1 |
|  | 563.8>892.5 | Qualifier | 35 | 19 | 0.90 | 0.71-1.1 |
| B | 687.4>841.5 | Quantifier | 35 | 18 | 1.25 | 1.11-1.45 |
|  | 687.4>766.4 | Qualifier | 35 | 23 | 1.25 | 1.11-1.45 |
|  | 687.4>865.5 | Qualifier | 35 | 23 | 1.25 | 1.11-1.45 |

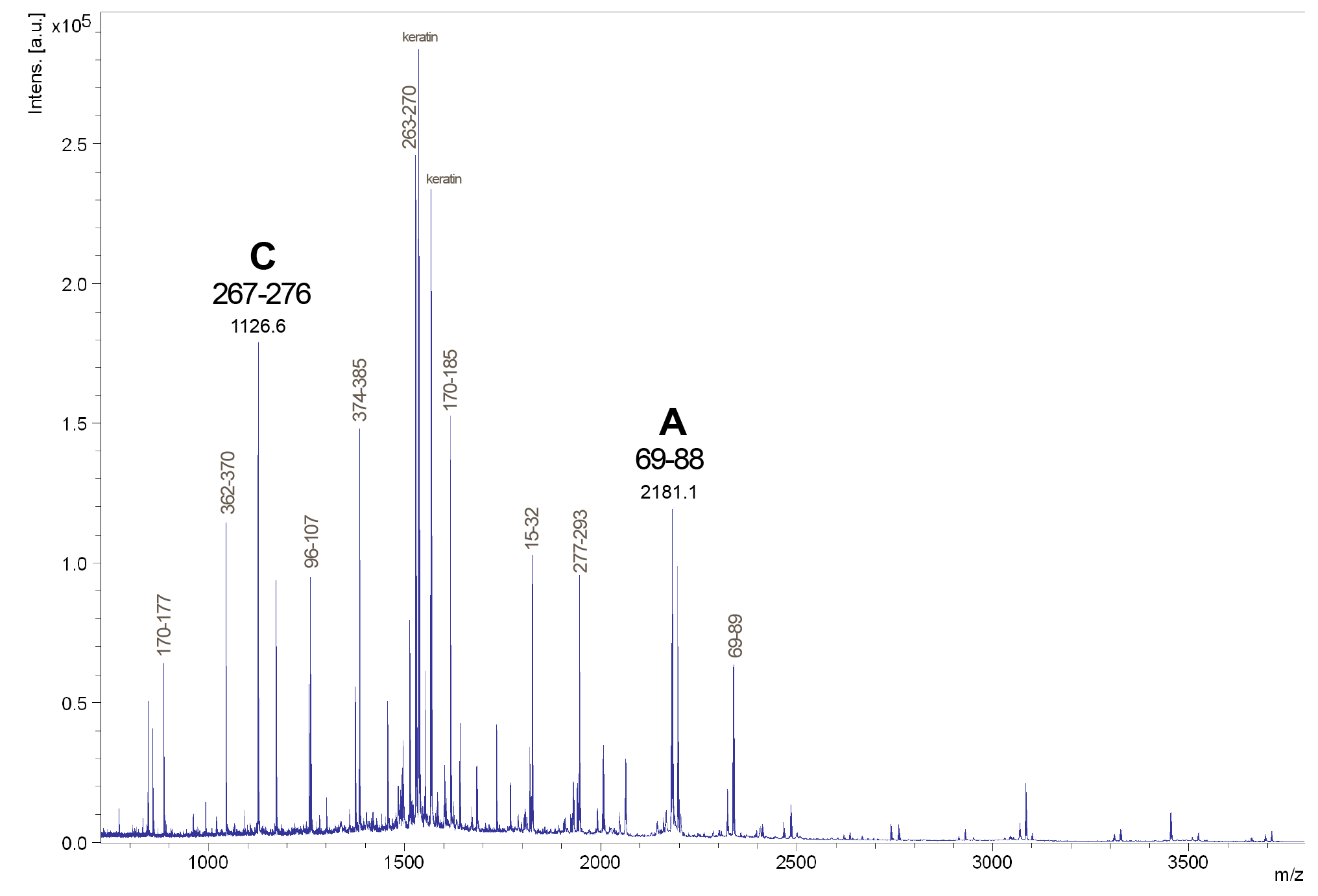

**Figure S1. MALDI-TOF mass spectrum of tryptic peptides from SARS-CoV-nucleoprotein**. Peptides from SARS-CoV-2 nucleoprotein are marked with their numbers in the amino acid sequence (see Figure 1). Peptides A and C are labeled.

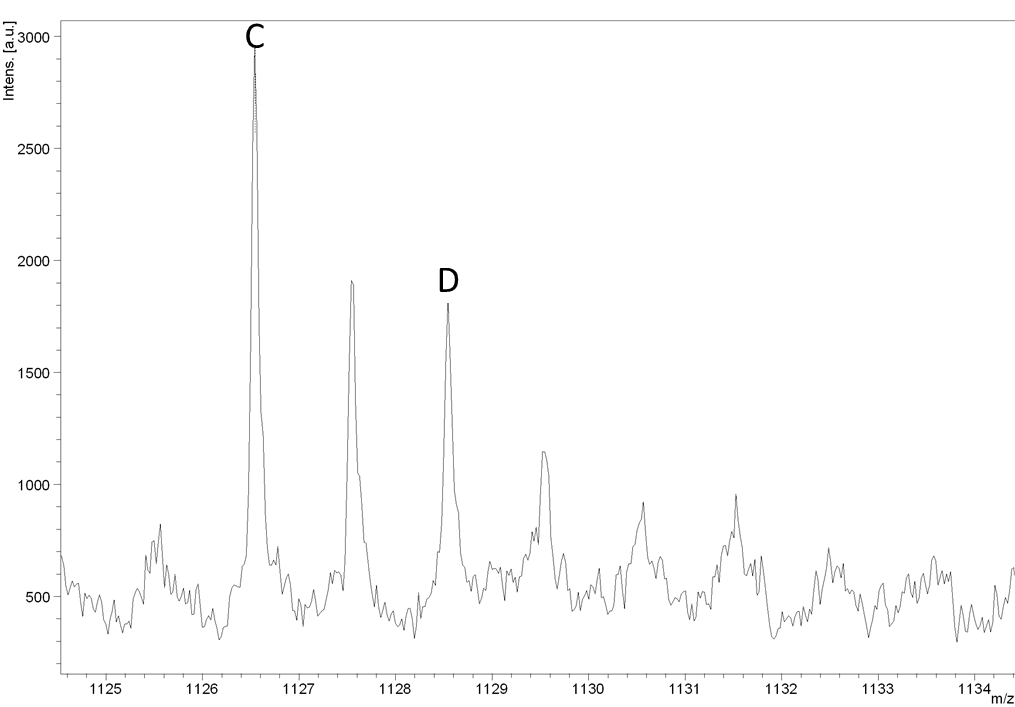

**Figure S2. MALDI-TOF mass spectrum of peptides C and D (antibody-enriched) from SARS-CoV-2 nucleoprotein.** Signals of peptide D (ADETQALPQR), [M+H]^+^ at *m/z* 1128.6 and peptide C (AYNVTQAFGR), [M+H]^+^ at *m/z* 1126.6 are shown enlarged.

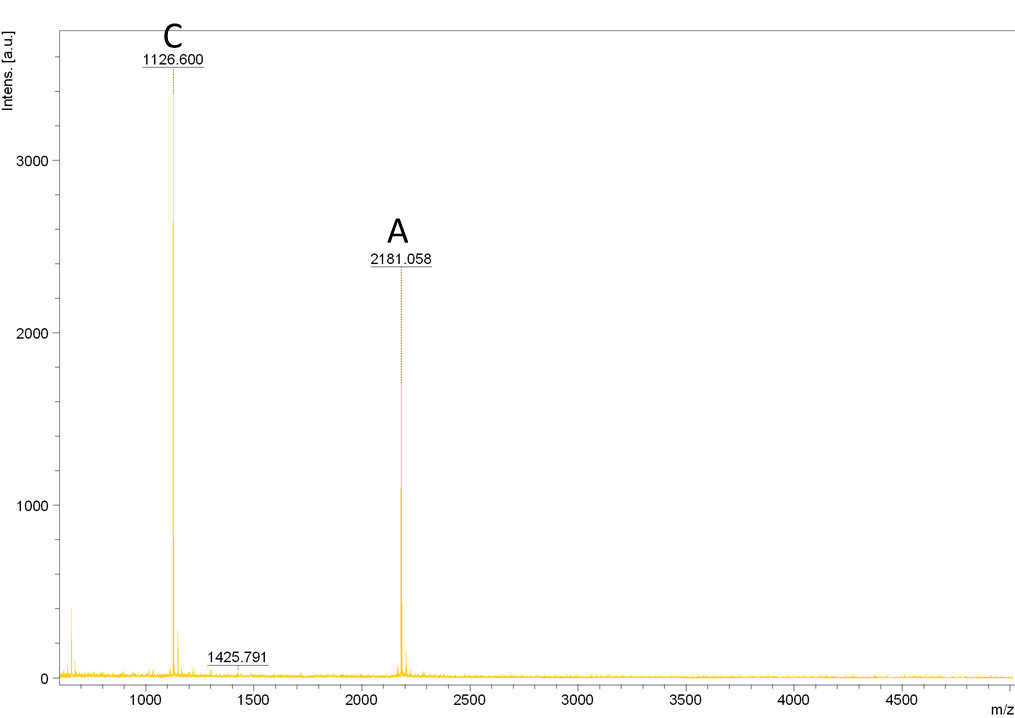

**Figure S3. MALDI-TOF mass spectrum of peptides A and C (antibody-enriched) from SARS-CoV-2 nucleoprotein, spiked into Cobas PCR Medium.** Peptides A and C are labeled.

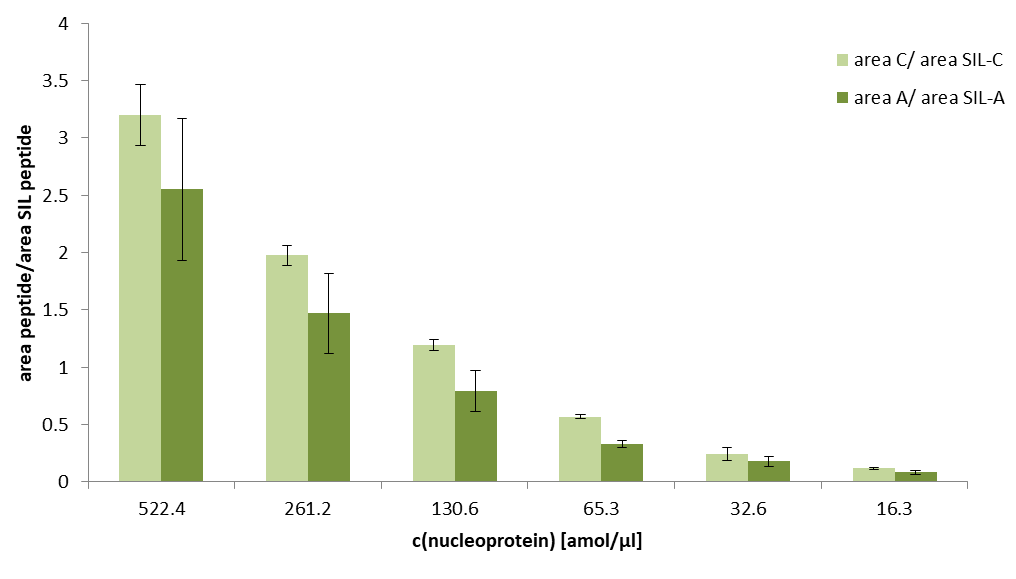

**Figure S4.** **Comparison of SARS-CoV-2 quantification with peptides SIL-A and SIL-C.** SARS-CoV-2 nucleoprotein was spiked at different concentrations (x-axis) into nasopharyngeal samples of healthy individuals (n=3). Samples were processed according to the workflow described in Figure 2 using MALDI-TOF-MS detection of peptides.

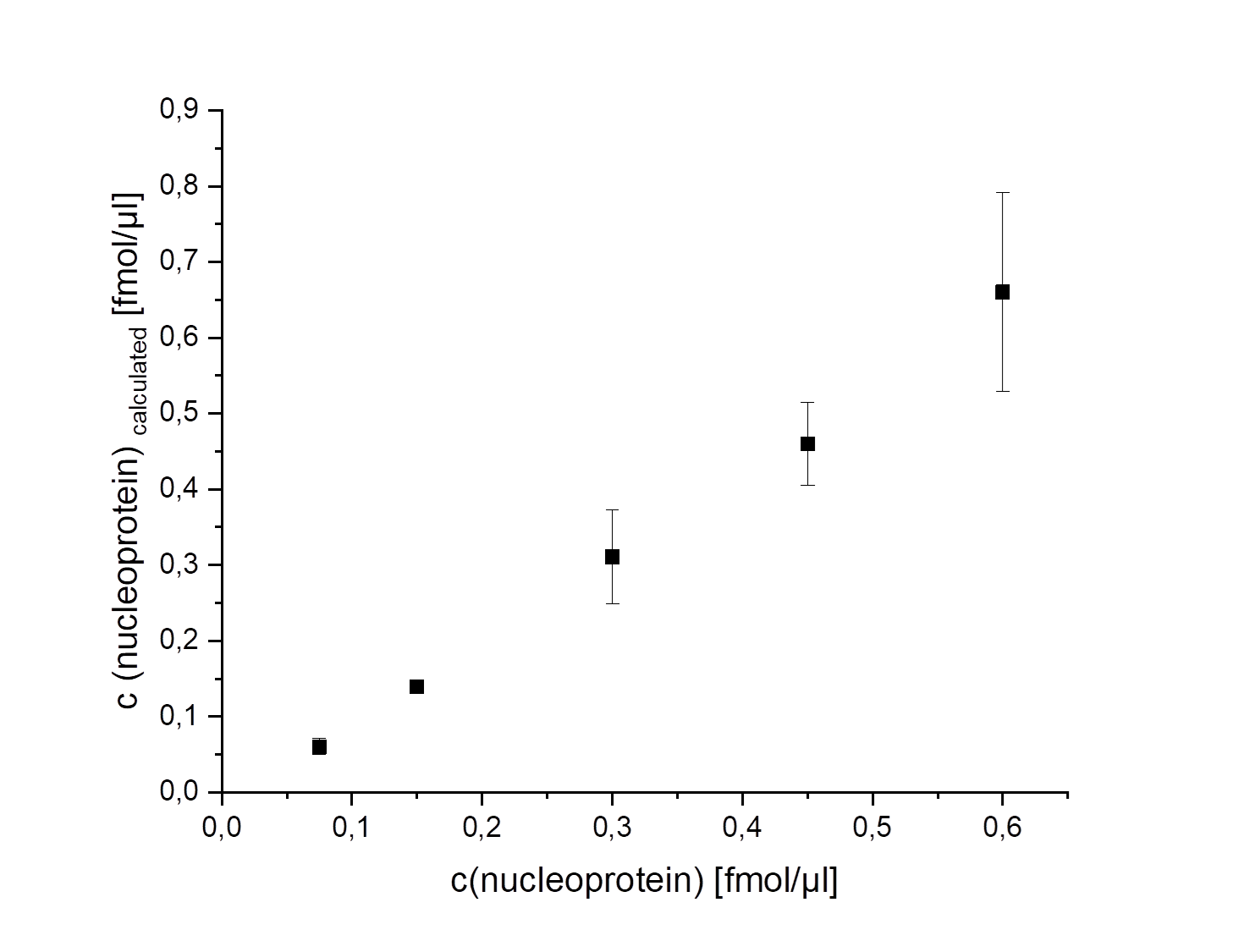

**Figure S5. Quantification of SARS-CoV-2 nucleoprotein with peptide SIL-C.** SARS-CoV-2 nucleoprotein was spiked at different concentrations (x-axis) into nasopharyngeal samples of healthy individuals (n=3). Samples were processed according to the workflow described in Figure 2 and concentrations (y-axis) were calculated by comparing the area of peptide C with the area of stable isotope-labeled peptide SIL-C.
